# Tracking the introduction and spread of SARS-CoV-2 in coastal Kenya

**DOI:** 10.1101/2020.10.05.20206730

**Authors:** George Githinji, Zaydah R. de Laurent, Khadija Said Mohammed, Donwilliams O. Omuoyo, Peter M. Macharia, John M. Morobe, Edward Otieno, Samson M. Kinyanjui, Ambrose Agweyu, Eric Maitha, Ben Kitole, Thani Suleiman, Mohamed Mwakinangu, John Nyambu, John Otieno, Barke Salim, Kadondi Kasera, John Kiiru, Rashid Aman, Edwine Barasa, George Warimwe, Philip Bejon, Benjamin Tsofa, Lynette Isabella Ochola-Oyier, D. James Nokes, Charles N. Agoti

## Abstract

We generated 274 SARS-CoV-2 genomes from samples collected during the early phase of the Kenyan pandemic. Phylogenetic analysis identified 8 global lineages and at least 76 independent SARS-CoV-2 introductions into Kenyan coast. The dominant B.1 lineage (European origin) accounted for 82.1% of the cases. Lineages A, B and B.4 were detected from screened individuals at the Kenya-Tanzania border or returning travellers but did not lead to established transmission. Though multiple lineages were introduced in coastal Kenya within three months following the initial confirmed case, none showed extensive local expansion other than cases characterised by lineage B.1, which accounted for 45 of the 76 introductions. We conclude that the international points of entry were important conduits of SARS-CoV-2 importations. We speculate that early public health responses prevented many introductions leading to established transmission, but nevertheless a few undetected introductions were sufficient to give rise to an established epidemic.

## Introduction

Severe acute respiratory syndrome coronavirus 2 (SARS-CoV-2), the aetiological agent of coronavirus disease 2019 (COVID-19), was first reported and confirmed in Kenya on the 13^th^ March 2020 (MoH, 2020b). This was shortly after the World Health Organisation (WHO) declared COVID-19 a pandemic on 11^th^ March 2020. SARS-CoV-2 outbreaks had previously been confirmed in many parts of Asia, Europe and North America following original emergence in Wuhan City, China, probably in December 2019 (Zhu et al., 2020). By end of June 2020, Kenya had reported 6,190 laboratory PCR confirmed SARS-CoV-2 cases and 144 COVID-19 associated deaths (MoH, 2020a). Within a month of the first confirmed case, Kenya put in place COVID-19 containment measures including closure of international borders, a dusk to dawn curfew, closure of all universities and schools, restaurants, bars and nightclubs, and religious meetings (churches, mosques and others). Meetings and social gatherings with more than 15 people were also banned. Movement into or out of areas that were considered epidemic hotspots became restricted, and the Government introduced strict quarantine procedures and isolation of infected individuals. Despite these public health measures, the number of new SARS-CoV-2 cases has increased steadily across the country and local transmission has now been established (MoH, 2020a; Ojal et al., 2020).

The Kenyan coast is an international tourism hub and a major gateway for the East and central Africa region. Although the government introduced screening at points of entry, SARS-CoV-2 infections may have been imported into the region by persons coming through the multiple points of entry and including by road from Nairobi (Figure 1A). The region has multiple ports of entry by air (Mombasa, Malindi and Lamu airports); land borders with Tanzania and Somalia; and a seaport in Mombasa (Figure 1B). Mombasa, Kenya’s second largest city, emerged as one of the epicentres of the early wave of SARS-CoV-2 infections (Ojal et al., 2020). Mombasa county has a population of over 1 million (2019 census) distributed across seven administrative sub-counties, and reported more infections than the other, more rural, Coastal Counties (i.e. Kwale, Taita Taveta, Kilifi, Tana River and Lamu). Within Mombasa county, Mvita island reported the largest number of cases (Figure 1C).

**Figure 1:**
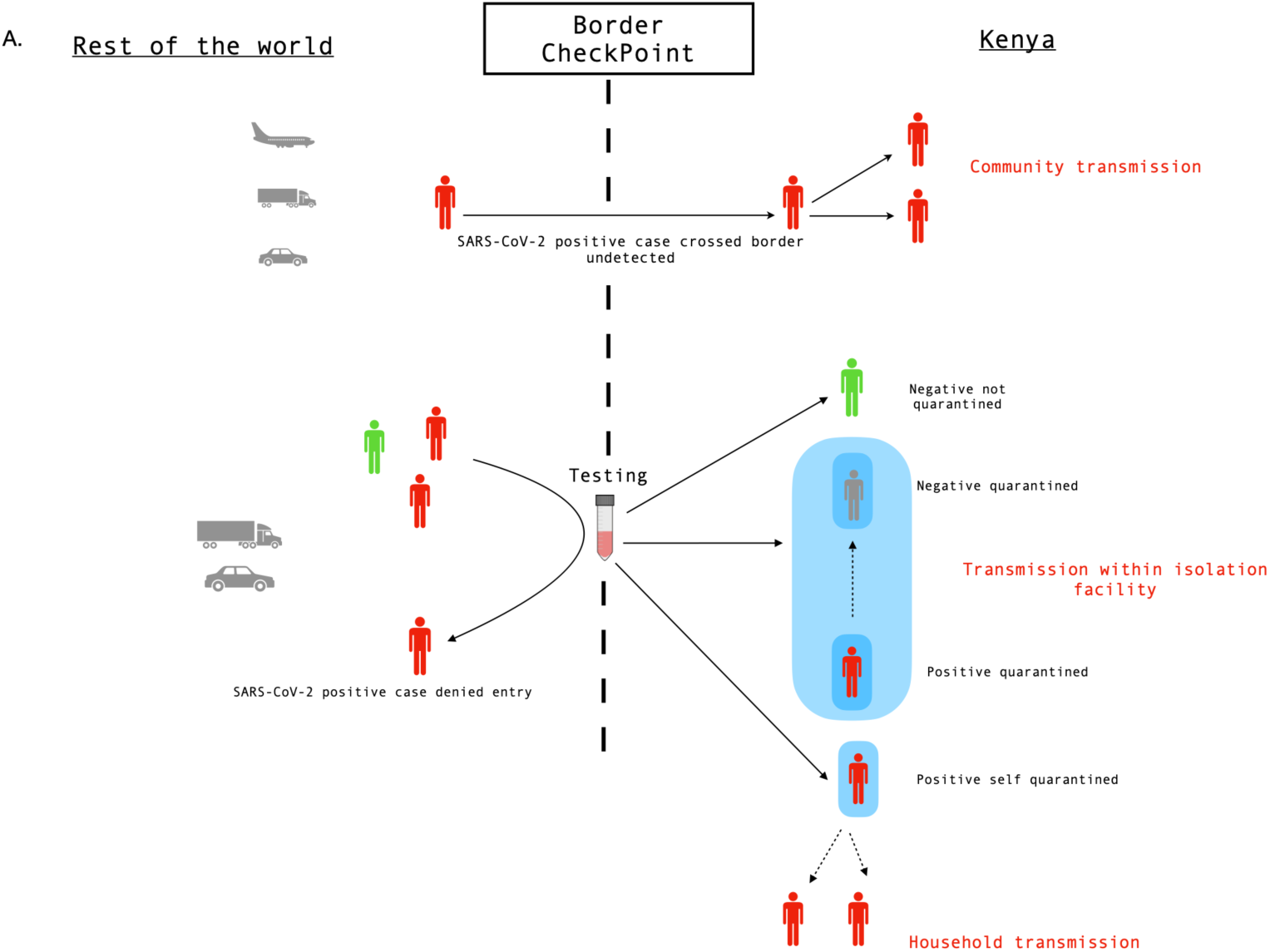
Importation model and geographical spread of SARS-CoV-2 at the Kenyan Coast: (A) SARS-CoV-2 cases are likely to have been imported into the region through international ports of entry undetected or through travellers or commercial truck drivers from regions like Nairobi and neighbouring countries. At the Tanzania border, Kenya implemented screening of commercial truck drivers and turned back individuals that were positive. Kenyan nationals were put in isolation facilities. Multiple undetected cases are likely to have been the source of the major epidemic that occurred in Mombasa. (B) A geographical map of Kenya showing the main administrative counties of Kwale, Mombasa, Taita Taveta, Kilifi, Tana River and Lamu and together comprise the coastal region. The total number of confirmed SARS-CoV-2 positive cases per hundred thousand across the coast as at of 24^th^ June 2020. The colour intensity is relative to the number of cases that were confirmed in the respective counties and overlaid on the major transportation infrastructure and hubs including road network, airport, seaport and border or international entry points. The cases detected in Taita Taveta were largely from the One Stop Border Post at Taveta/Holili crossing point between Southern Kenya and Northern Tanzania. (C) A map of Mombasa county showing the distribution of the number of confirmed SARS-CoV-2 positive at the sub-county administrative level. One-hundred and twenty-two cases that did not have sub-county information and are not shown.

Information on the early importation and spread of SARS-CoV-2 in Kenya is important in assessing the effectiveness of the early interventions and for designing additional COVID-19 control measures in Kenya and the East Africa region. Similar investigations in Iceland, Singapore and Europe have been undertaken, but to date there are few datasets from Africa and none with large numbers of isolates from a single region to show the characteristics of the early spread of SARS-CoV-2. We sequenced and phylogenetically analysed 274 PCR positive cases detected between March and June 2020 on the Kenyan Coast.

## Results

### SARS-CoV-2 testing and sequencing on the Kenyan Coast

SARS-CoV-2 RT-PCR testing was set up at KEMRI-Wellcome Trust Research Programme in Kilifi (KWTRP) in mid-March 2020. This became the government designated testing centre to support the Department of Public Health Rapid Response Teams (RRTs) from the six coastal counties of Kilifi, Kwale, Lamu, Mombasa, Taita Taveta and Tana River (Figure 1). The testing criteria was dependant on the guidelines from the ministry of health and could be divided into roughly four phases (Table S2) depending on the number of cases that were reported in the country. By June 30^th^, 1,257 out of 25,492 samples tested were confirmed RT-PCR positive across five of the six coastal counties namely Mombasa (n=973), Kwale (n=129), Taita Taveta (n=109), Kilifi (n=44) and Lamu (n=2) (Figure 1 and Figure 2). Between March and 24^th^ June 2020, RRTs obtained samples from repatriated citizens including among individuals arriving at ports of entry and from persons presenting at major hospitals with symptoms consistent with SARS-CoV-2 infection, contacts of confirmed cases and mass testing of residents in Mombasa. By June 30^th^, 108 and 79 tests were carried out in Lamu and Tana River counties respectively. Only two positive samples were identified in Lamu, reported on the 27-June-2020. No positive samples had been detected in Tana River county by end of June 2020 (Figure 2).

**Figure 2:**
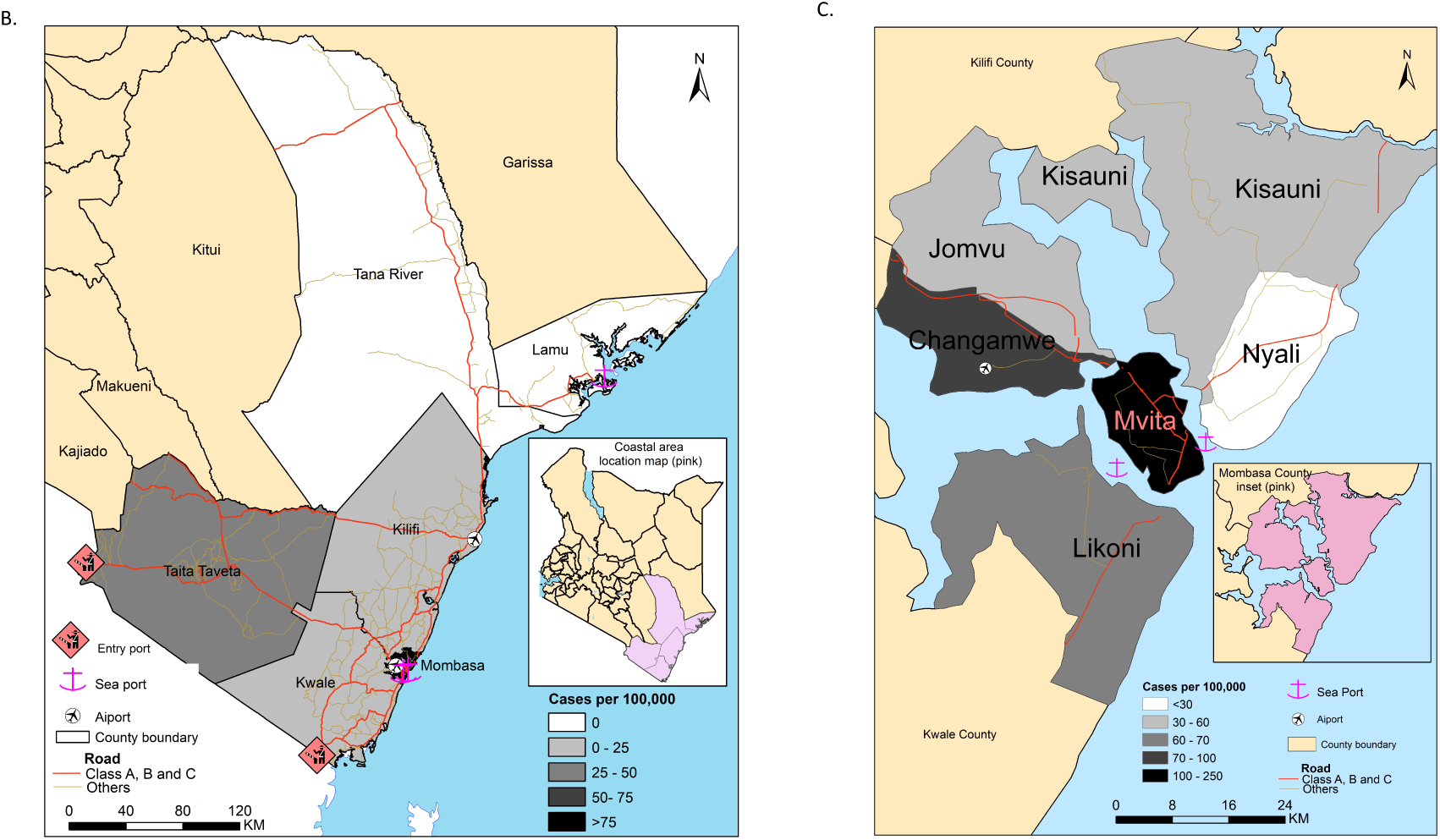
Testing of SARS-CoV-2 cases at the Kenya Coast: (A) The cumulative number of SARS-CoV-2 tests (grey line) and the number of positive cases per week (brown line) over the study period. Thee major public health measures undertaken by the Kenyan government during the study period are represented by the horizontal bars and shaded based on the length of the intervention. (B) The cumulative number of SARS-CoV-2 positives cases that were confirmed from each of the four counties at the Kenyan coast are represented by the lines. The horizontal bars represent the county specific public health interventions that were undertaken during the study period and early in the epidemic period. The length of the bars corresponds to the time duration for each respective intervention.

We analysed sequence data from 274 SARS-CoV-2 RT-PCR positive samples collected between 17^th^ March 2020 and 24^th^ June 2020 representing majority of cases identified in our laboratory (Figure 2). The 274 genomes came from four of the six coastal counties (Mombasa (212), Kwale (n=32), Taita Taveta (22), and Kilifi (8)) and provided sequence data that was more than eighty percent complete. This included follow-up samples from 19 individuals (Figure S1). The median age of the individuals from the sequenced samples across all the counties was 39 years (range 0 to 85 years), 62% of whom were male, and half were asymptomatic (Table 1 and Table S1). A number of the sequenced samples were from persons (20.4%) with a history of recent travel (i.e. international travel within last two weeks) or were sampled (19.3%) at the port of entry into Kenya (Table 1 and supplementary Table S1).

**Table 1:**
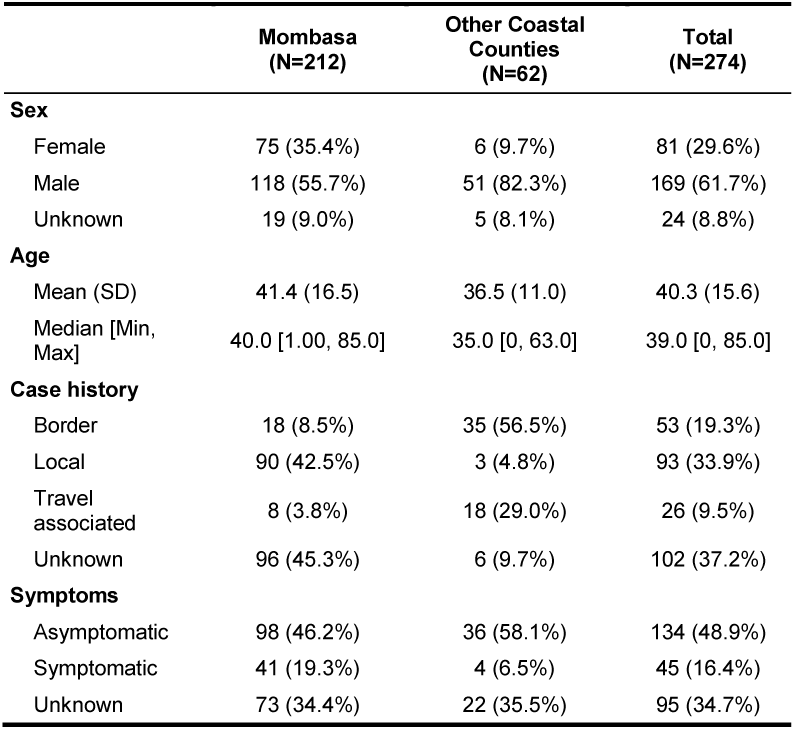
Demographic characteristics of SARS-CoV-2 whole genome sequenced samples (>80% complete) collected between March and June 2020 (n=274) from coastal Kenya stratified by county. The case history demographic characteristic was derived from both self-reported travel history and presentation at a border point. Local case-history refers to individuals that did not report a history of travel and neither were they screened at a port of entry. Individuals whose case histories were not filled, or information was missing were labelled as unknown. (See also Table S2)

|  | **Mombasa**<br>**(N=212)** | **Other Coastal**<br>**Counties**<br>**(N=62)** | **Total**<br>**(N=274)** |
| --- | --- | --- | --- |
| **Sex** |  |  |  |
| Female | 75 (35.4%) | 6 (9.7%) | 81 (29.6%) |
| Male | 118 (55.7%) | 51 (82.3%) | 169 (61.7%) |
| Unknown | 19 (9.0%) | 5 (8.1%) | 24 (8.8%) |
| **Age** |  |  |  |
| Mean (SD) | 41.4 (16.5) | 36.5 (11.0) | 40.3 (15.6) |
| Median [Min, Max] | 40.0 [1.00, 85.0] | 35.0 [0, 63.0] | 39.0 [0, 85.0] |
| **Case history** |  |  |  |
| Border | 18 (8.5%) | 35 (56.5%) | 53 (19.3%) |
| Local | 90 (42.5%) | 3 (4.8%) | 93 (33.9%) |
| Travel associated | 8 (3.8%) | 18 (29.0%) | 26 (9.5%) |
| Unknown | 96 (45.3%) | 6 (9.7%) | 102 (37.2%) |
| **Symptoms** |  |  |  |
| Asymptomatic | 98 (46.2%) | 36 (58.1%) | 134 (48.9%) |
| Symptomatic | 41 (19.3%) | 4 (6.5%) | 45 (16.4%) |
| Unknown | 73 (34.4%) | 22 (35.5%) | 95 (34.7%) |

### Phylogenetic clustering of sequences from coastal Kenya

We estimated the number of independent introductions of SARS-CoV-2 into coastal Kenya by phylogenetic analysis of the local sequences against a background of 983 genomes sampled globally (December 2019-June 2020). We examined pairwise nucleotide differences and examined the travel history of the sequenced cases. From the maximum-likelihood phylogenetic tree, we observed that the coastal Kenya genomes were distributed across multiple clades in the global tree (Supplementary figure 1). The phylogenetic interspersing of the local viruses within the global sample provided evidence of multiple viral introductions into the local population (Figure 3 and Supplementary figure 1), resulting in 76 independent SARS-CoV-2 introductions into coastal Kenya from this analysis (Table 2). The observed local clusters were varied in both size and diversity, and some were suspected to have comprised further multiple independent introductions of closely related viral lineages. The results from the temporal clustering (Figure 4) were consistent with known case history and with our expectations of SARS-CoV-2 importations into the region (Figure 1A).

**Table 2:** A summary table of early SARS-CoV-2 introductions into the Kenyan Coast stratified by lineage. Each row represents an observed lineage, and the corresponding number of introductions in each lineage. The third column represents the first-time an introduction from each of the lineages was observed and entry route into the region and whether the introductory case was associated with COVID-19-like symptoms at the time of sampling.

| Lineage | Approximate | Proportion of | Date of Initial | Entry Source | Symptoms | Comments |
| --- | --- | --- | --- | --- | --- | --- |

|  | Introductions | sequences | Case |  |  |  |
| --- | --- | --- | --- | --- | --- | --- |
| **A** | 7 | 2.9 | 05-Apr-2020 | Household/After entry point | Yes | Returning from Dubai |
| **B** | 2 | 0.72 | 19-Apr-2020 | Household/After entry point | No | Returning from Dubai |
| **B.1** | 45 | 82.1 | 17-Mar-2020 | Local/surveillance | Yes | History of travel |
| **B.1.1** | 14 | 11.31 | 02-April-2020 | Local/surveillance | Yes | No history of travel |
| **B.1.1.1** | 1 | 0.36 | 16-May-2020 | Border/Entry | No | Travel history to Tanzania/Tanzanian National |
| **B.1.1.33** | 1 | 0.36 | 13-May-2020 | Mass testing | No | Detected during mass testing |
| **B.1.5** | 3 | 1.09 | 19-Apr-2020 | Household/After entry point | No | Travel history |
| **B.4** | 3 | 1.09 | 05-Apr-2020 | Border/ Entry point | No | Two sampled at port of entry; 1 travelled to Zambia |

**Figure 3:**
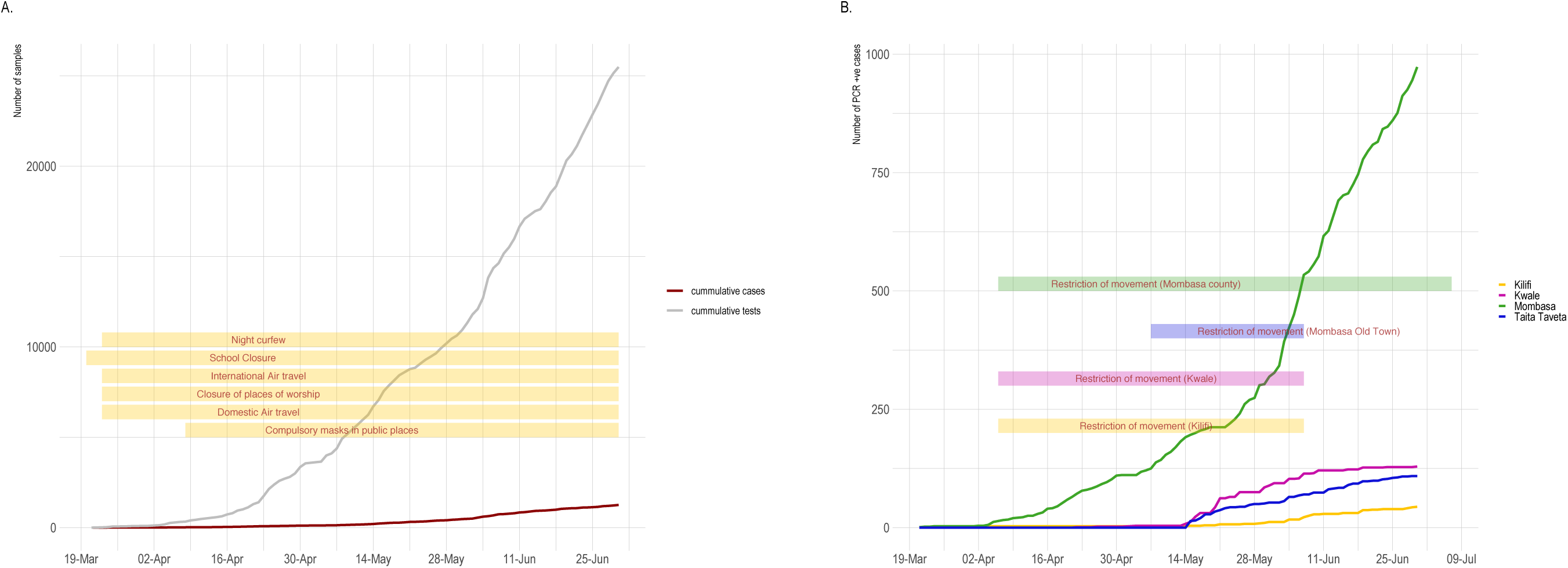
Sequence and genetic diversity of sequences during the early phase of the epidemic in coastal Kenya: (A) A bar graph showing the proportion of assigned lineages to 274 SARS-CoV-2 sequences collected from the coast between 24^th^ March to 24^th^ June 2020 and stratified by county. The colours represent the 8 main lineages that were identified. (B) A bar graph showing the proportion of 274 SARS-CoV-2 sequences from the coastal region and the associated overall epidemiological source information aggregated by week and stratified by county. The colours represent the main epidemiological sources (border, local or travel associated) that were associated with the infections.

**Figure 4:**
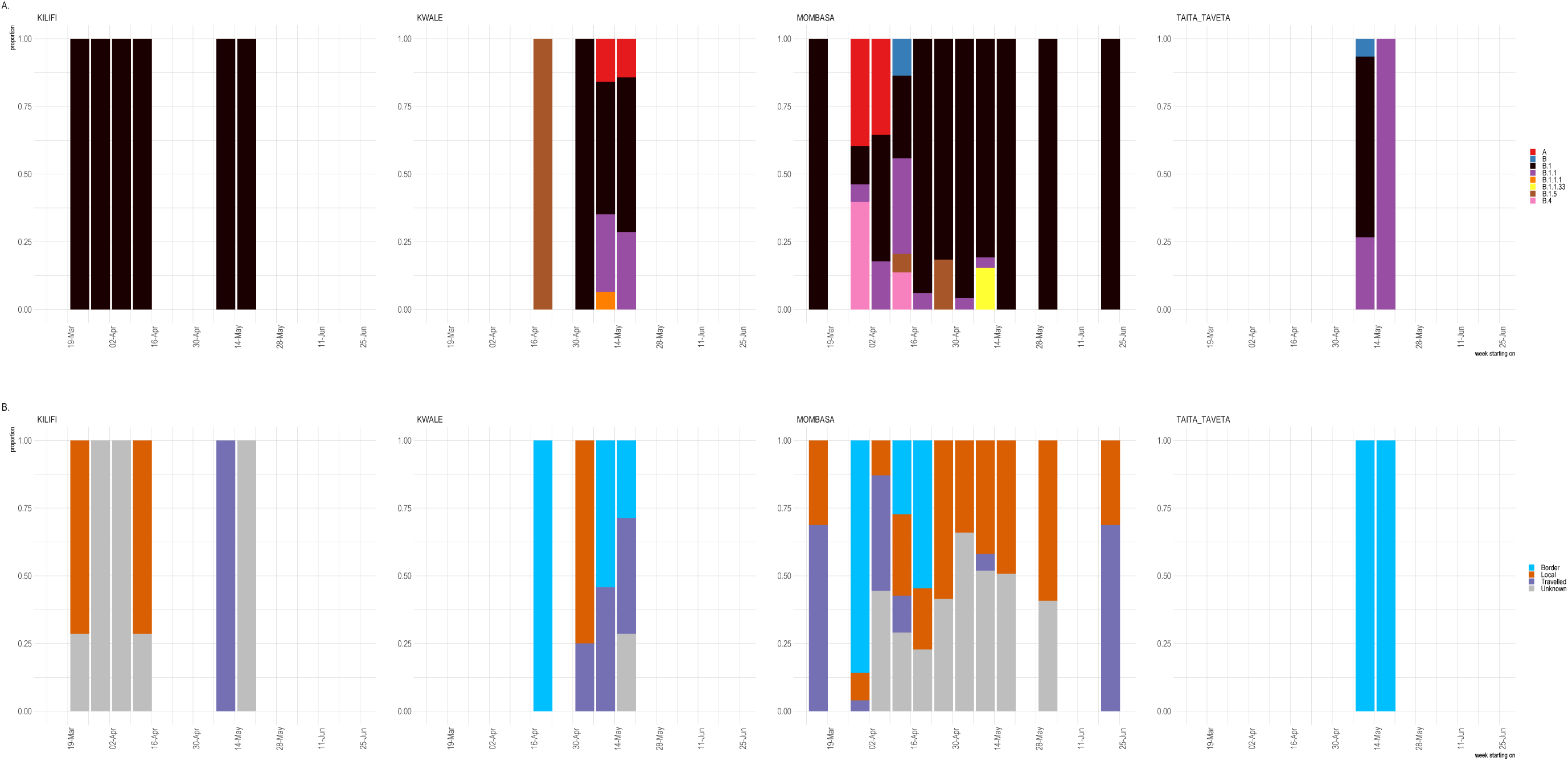
A time resolved Bayesian phylogenetic tree of sequences collected from the coast Kenya stratified by: (A) Geographical location. The tip colour represents samples collected from each of the coastal counties (B) A time tree showing the PANGOLIN assigned lineage for each of the coastal sequences (C) A time tree showing the samples stratified based on the travel history and detection at the border point. Local sequences were individuals with no history of travel.

## Introduction of global circulating lineages into coastal Kenya

We used a dynamic lineage classification method to identify relevant epidemiologically important lineages using the Phylogenetic Assignment of named Global Outbreak LINeages (PANGOLIN) v2.0.4 toolkit (Rambaut et al., 2020b) to classify the Kenyan sequences into 8 lineages (Figure 3A). Most of the sequences (93.8%) were assigned into the lineages with a probability of greater than 0.7. The dominant lineage at the coastal region was B.1, which was first sampled in March and accounted for 82.1% of the sequenced cases (n=225). Based on global data, the B.1. lineage comprised the majority of the large Italian and UK outbreaks and was observed in several other European outbreaks and has spread to the rest of the world. (Rambaut et al., 2020b). In coastal Kenya, B.1. was the only detectable lineage among the early cases observed in March and continued to dominate the cases in the coastal region including the most recent samples sequenced from the month of June 2020 (n=5) (Figure 3). Based on self-reported history of travel, approximately 40% of infected individuals had no history of travel outside their localities (Table 1, Figure 4C) providing evidence for established local transmission. Overall, we observed multiple lineages across the coastal counties (Mombasa (n=7), Kwale (n=5) and Taita Taveta (n=3) other than in Kilifi (n=1). Mombasa, Kwale and Taita Taveta have multiple ports of entry into Kenya (Figure 1B and C). The B.1 lineage was the dominant lineage across coastal counties except for Taita Taveta which was dominated by the B.1.1, albeit from among only 22 cases (Figure 4B). Interestingly, the sequenced cases from Taita Taveta were from male individuals aged between 20 and 60 years and were all detected at the border point, suggesting that these infections may have been acquired outside Kenya (supplementary Table S1).

The B.1.1 lineage was the second most observed in our sequenced sample set (n=31, 11.3%) and was repeatedly observed among individuals sampled at the ports of entry (n=17) (Figure 3, Table 2 and 2) or those with a history of travel (n=3). This lineage is defined by three single nucleotide polymorphisms (SNPs) at positions G28881A, G28882A, G28883C of the genome. These sequences were from individuals admitted to isolation centres which may partly explain the lineage’s limited subsequent local expansion. Both phylogenetic analysis (Figure 4) and pairwise sequence comparison suggest that this lineage comprises of two increasingly diverging clusters (Figure 6B).

We observed 8 lineage A associated sequences in Mombasa and Kwale between April and May 2020. These sequences were associated with individuals that presented at the Kenya/Tanzania border point (Table 1 and Table 2). A separate introduction of lineage A was observed from an individual who had travelled from Dubai (Table 2). Lineage A is characterised by the early Chinese sequences (Rambaut et al., 2020a). However, following these introductions there was no evidence of ongoing transmission from lineage A viruses.

The lineages B (n=2), B.1.1.1 (n=1), B.1.1.33 (n=1), and B.1.5 (n=1) and B.4 (n=3) were observed less frequently than lineage B.1. The 3 lineage B.4 cases were sampled from Mombasa from two individuals that were screened at a point of entry (one of who had a travel history from Lusaka, Zambia) and one individual who had no travel history. The B.4 lineage has been previously reported from individuals who had a travel history from Iran (Rambaut et al., 2020b).

A majority of the sequences were characterised by between 4 and 7 nucleotide differences relative to the Wuhan reference sequence (NC_045512.2) (supplementary figure 2). Sequence to sequence pairwise divergence was observed across all sequences and within lineages (Supplementary figure 2B). Sequences corresponding to lineage B.1 were associated with more pairwise differences, while those corresponding to B.1.1 showed a bimodal distribution of sequence diversity from the sequence information (Supplementary figure 2). Taken together, these observations support multiple importations into the Kenyan coast because it is unlikely that the observed differences were a result of few or a single introduction. Given the dominance of B.1 lineage during the early phase of the epidemic, it is not surprising that many of the coastal sequences contained the spike D614G variant (Figure 6C). This mutation arose early during the epidemic (Korber et al., 2020) and now dominates the majority of the cases across the globe (Volz et al., 2020).

A few sequences were collected at multiple timepoints from 19 individuals who were placed in quarantine facilities (supplementary figure 3). These individuals were infected by SARS-CoV-2 viruses that corresponded with the B.1 (n=17), A (n=1) and B.1.1 (n=1) lineages. Individual R_5 was infected by a B.1 lineage however the sample collected at the last timepoint was assigned a B lineage, raising the possibility of cross-infection during quarantine. Similarly, individual R_10 was infected by a lineage B.1 but the sequence from the last timepoint was assigned lineage B.1.1 (supplementary figure 3).

## Discussion

In this work we used genomic surveillance of SARS-CoV-2 to elucidate the origins and transmission of SARS-CoV-2 among the Kenyan coastal population. Our genomic sequence and epidemiological data reveal at least 76 independent introductions of SARS-CoV-2 in the Coastal region of Kenya. We saw evidence of local transmission of the B1 lineage in the Coastal region particularly in Mombasa. Although several major global viral lineages were detected in returning citizens or international travellers, at the entry points or within the country, few resulted in large outbreaks (Figure 4 and supplementary figure 1). A possible explanation for this is that the government actions of enhanced border control and screening, quarantine and isolation of positive cases were effective in mitigating transmission. In addition, restrictions in international air-travel into the country reduced the number of potential introductions. This is corroborated by data showing low R0 values in the early phase of the epidemic (Ojal et al., 2020). Differences in the transmissibility of the different lineages is possible (Korber et al., 2020; Lorenzo-Redondo et al., 2020) but in our case the predominance of B1 in local transmission can be explained by the fact that most introductions were of B.1 lineages. The detection of a unique lineage (B.1.1.33) in a sample that was collected during mass testing (13-May-2020) in the Mombasa City could be indicative of cryptic transmission within the community in spite of the intervention, but also could be failure to detect this lineage at the port of entry due to sampling limitations.

Early COVID-19 control strategies by the Kenyan government were geared towards preventing establishment of community transmission. These policies appear to have been largely successful in that most of the introductions were not associated with subsequent established transmission. Nevertheless, a minority of introductions did go on to establish sustained transmission and give rise to local transmission despite an absence of new virus lineages being imported. This underlines the severe challenge to a strategy aimed at preventing the introduction of virus as any cases escaping the net can potentially establish community spread.

A number of samples (n=35) that were analysed in this study were collected at the Kenya-Tanzania border points. In neighbouring Uganda, international truck drivers including those from Kenya were identified as common sources of the infection (Bugembe et al., 2020; Nakkazi, 2020). This led to mass testing of truck drivers in Kenya and a requirement of a “COVID-19 free certificate” before leaving or entering Kenya. The genomic analysis of SARS-CoV-2 cases in Uganda detected lineages similar to those reported in our study (i.e. A, B, B.1, B.1.1, B.1.1.1 and B.4). This finding emphasizes the need for common strategies across East Africa to effectively control the pandemic. Furthermore lineage B.1 dominance in coastal Kenya was consistent with what has been reported in some parts of the world including several African countries (TIBA, 2020). Globally, the B.1 lineage was a major European lineage first identified on February 15^th^, 2020. It will be interesting to continue evaluating the local spread and sustainability of this lineage in Africa as the epidemic evolves.

A limitation of our analysis is the incomplete genomes due to amplicon dropouts and which resulted in analysis of fewer genomes. In addition, the collection of epidemiological data was partially complete for a number of demographic characteristics (Table 1, supplementary table S1). The testing strategy was not systematic, and guidelines were repeatedly revised as the epidemic evolved (supplementary table 2). This might have created a bias in the observed lineage prevalence. Although we present a substantial number of genomes from a single region within East-Africa, these data may not generalize across the region. Nevertheless, our data provides a background for monitoring the progress of the SARS-CoV-2 epidemic in Kenya and a platform for continued surveillance and to build evidence for routes of transmission, sustained spread, effectiveness of control interventions and evolution of the virus within the local communities.

## Conclusion

Overall our data provides evidence for both limited spread of lineages that came in through the border. Screening at the border and early surveillance efforts with contact tracing or isolation and quarantine of identified cases appears to have been successful in preventing the majority of introductions leading to further transmission. Nevertheless, a number of cases went undetected (Figure1A) and led to community transmission. This is particularly clear when looking at the cases attributed to lineage B.1 in Mombasa (Figure 3 and Figure 4).

Our analysis revealed the extent of imported SARS-CoV-2 genomic diversity that was seeded in local communities and the corresponding routes of novel introductions. Most of the introductions detected at border points did not result in community transmission, which reinforces the importance of SARS-CoV-2 testing at border crossing points and strict quarantining of positive cases during the early phases of an epidemic when the goal was to prevent transmission becoming established, but also the fragility of such methods when a few cases remain undetected. These data have assisted to evaluate the effectiveness of recent government public health interventions and were rapidly shared with the Kenya Ministry of Health to inform on relevant public health response.

## Methods

### Ethics statement

The samples were collected under the Kenya Ministry of Health (MoH) protocols as part of the national response to the COVID-19 pandemic. The whole genome sequencing study protocol was reviewed and approved by the Scientific and Ethics Review Committee (SERU) that sits at the Kenya Medical Research Institute (KEMRI) headquarters in Nairobi (SERU # 4035).

### Study site and population

The sequenced specimens were collected between 17th March to 24^th^ June 2020 by the Rapid Response Teams (RRTs) from four counties (Kilifi, Mombasa, Kwale, and Taita Taveta). The eligibility criteria for SARS-CoV-2 testing was as provided by the Ministry of Health guidelines during the period covered in this analysis (Supplementary text S1 and supplementary table S2).

### SARS-CoV-2 diagnosis

Nasopharyngeal (NP) and oropharyngeal (OP) swabs were collected into a single tube and transported to KEMRI-Wellcome Trust Research Programme (KWTRP) for SARS-CoV-2 diagnosis. Briefly, the positive infections were RNA purified using a QIAGEN extraction kit followed by real-time reverse-transcription PCR (RT-PCR) (Agoti et al., 2020; Said et al., 2020).

### SARS-CoV-2 genome amplification, library preparation and sequencing

Positive SARS-Cov-2 samples were sequenced using a modified tilled-amplicon PCR approach based on the ARTIC V2 protocol and V2 and V3 primer sets. Sequencing libraries were prepared and sequenced using ONT version 9.4.1 flow-cells. Consensus sequences were assembled using the ARTIC bioinformatics protocol from base-called and demultiplexed FASTQ reads as described in the detailed methods section (text S1). Detailed quality control of the resulting genomes was done using manual and automated methods. Detailed analysis of mutations was conducted and visualised using Webclades (v0.4.2). Viral lineages were assigned using Pangolin (v2) toolkit.

### Sampling from global sequence dataset

A total of 62,644 SARS-CoV-2 whole genome sequences were downloaded from the Global Initiative on Sharing All Influenza Data (GISAID) sequence database (https://www.gisaid.org/) at a single time point. The sequences were filtered to remove erroneous, incomplete and improperly formatted data. Sequences shorter than 29,500 nucleotides those that were not accompanied by the day and month of collection were excluded. All sequences collected later than 24^th^ June 2020 were also excluded. The resulting dataset was then grouped based region, country and pangolin lineage and two sequences were picked to represent each lineage from each country. The remaining 1,439 sequences were then subjected to a quality check based on NextClade quality filters and visual inspection. The sequences that passed this quality filter (n=983) were referred to as the global dataset and were used to place the Coastal Kenya sequences within a global phylogenetic context.

### Sequence alignment and phylogenetics clustering

Multiple sequence alignments were done using MAFFT v7.310. The alignments were masked and edited to remove untranslated regions and known problematic regions. To place SARS-CoV-2 sequences collected from the coast within a global content, model selection was conducted and inferred using the IQTREE (version 2.0.3) model selection tool and a maximum likelihood phylogenetic tree were estimated using the GTR+G+R3 substitution model. To estimate evolutionary relationships between sequences collected in coastal Kenya, a separate maximum likelihood phylogenetic tree was inferred from an alignment of 274 coastal Kenya sequences in addition to the NC_045512.2 reference sequence using RAxML-NGS v0.9.0) and run with 1000 bootstraps. The tree was rooted using the reference sequence as an outgroup. A time-scaled phylogeny was generated using the BEAST (v1.10.4) Bayesian phylogenetic framework to reconstruct a time-scaled phylogeny from 274 sequences from the Kenyan coast. The BEAST analysis was parameterized with an uncorrelated log-normal relaxed-clock (UCLN) model with an exponential growth coalescent prior and the HKY85 substitution model. Four independent Markov chain Monte Carlo (MCMC) runs of 100 million generations were performed with sampling every 10,000 generations. MCMC convergence and log files from the runs were inspected using Tracer v1.7 (reference). A maximum clade credibility (MCC) tree was generated using TreeAnnotator version (references).

### Estimating the number of importations of SARS-CoV-2 viruses at the coast

We estimated the minimum number of importations by triangulating between the known number of travel cases and manual inspection of the phylogenetic trees corresponding to each lineage and the global phylogenetic tree.

## Data Availability

SARS-CoV-2 sequence data used in this analysis are publicly available from GSAID
Source code that was used in the analysis of this work is available from GitHub (https://github.com/george-githinji/SARS-CoV-2_KE) and data used to create the figures can also be found in the supplemental files.

## List of Figures

**Supplementary figure 1: Phylogenetic clustering:**

(A) A maximum likelihood phylogenetic tree showing the evolutionary relationship between whole genome sequences SARS-CoV-2 cases collected from the coastal counties of Kenya (n=274) against a background of 983 global sequences shown in grey tips.

**Supplementary figure 2:**

A) A scatter plot showing the number of mutations acquired by sequences collected from the coast relative to the reference genome (x-axis) and the number of ambiguous nucleotides in the sequences (y-axis).

B) A Violin plot showing the number of pairwise nucleotide differences (y-axis) distribution within the 274 SARS-CoV-2 sequences and between sequences of different lineages (x-axis).

(C) The relative proportion (y-axis) of known mutations (x-axis) among the sequences collected from the Kenyan coast.

**Supplementary figure 3**:

(A) Temporal shedding patterns. Individuals that were taken to an isolation facility provided multiple samples some of which were sequenced. The figure shows samples taken at multiple time points from the same individual and coloured by predicted Pangolin lineage.

(B) Each panel shows a highlighter plot representing the position of nucleotide differences (x-axis) from each set of sequences (y-axis) taken from the same individual. Grey regions represent missing sequence data.

## Lead contact

Further information and requests for resources and reagents should be directed to and will be fulfilled by the Lead Contact, George Githinji

## Materials Availability

This study did not generate new unique reagents, but raw data and code generated as part of this research can be found in the Supplemental files as well as on public resources as specified in the Data and Code Availability section below.

## Data and Code Availability

- SARS-CoV-2 sequence data used in this analysis are publicly available from GSAID
- Source code that was used in the analysis of this work is available from GitHub (https://github.com/george-githinji/SARS-CoV-2_KE) and data used to create the figures can also be found in the supplemental files.

## METHOD DETAILS

### Study site and population

The sequenced specimens were collected by Rapid Response Teams (RRTs) in four counties of Coastal Kenya (Kilifi, Mombasa, Kwale, and Taita Taveta) between 17th March to 24^th^ June 2020. The eligibility criteria for an individual to be tested for SARS-CoV-2 was revised by the Ministry of Health during the period covered in this analysis. For ease of presentation, we split the analysis period into four phases (supplementary table S2). During the first phase, individuals were eligible for testing if they showed specific respiratory illness symptoms (cough, fever or difficulty in breathing) and had a recent history of international travel or were a listed as close contact of a confirmed case. Returning travellers from early affected countries (China, Iran, Italy and USA) were of most interest during phase 1. Phase two began after the first SARS-CoV-2 confirmed case in Kenya (12^th^ March 2020). In addition to guidelines that were provided during phase 1, the government ordered that everyone arriving by air from international visits was to proceed to a quarantine facility for two weeks. At the end of the quarantine period, an individual was release after a negative RT-PCR test. Phase 3 began when the government suspected that community transmission was ongoing in some parts of Mombasa and Nairobi city based on increased number of positive cases from these regions. The government recommended and rolled out mass testing of workers at the Kenya Ports Authority (KPA) in Mombasa and the general public in the Mvita sub-county (also referred to as the Old town or Mombasa island) (Figure 1). Phase 4 began after reports that international truck drivers played a role in transmission of the virus which was also noted by border entry surveillance teams in land-locked Uganda. The Kenya Ministry of Health (MoH) ordered mass testing for truck drivers including those entering Kenya through the Lunga Lunga border (Kwale county) and Holili port (Taita-Taveta) county. Our laboratory testing efforts throughout this period included patients with respiratory symptoms consistent with COVID-19 turning-up at the Aga Khan Hospital, Mombasa Hospital, Coast Provincial Referral and Teaching Hospital (CPRTH) and The Premier Hospital.

### SARS-CoV-2 diagnosis at KWTRP

The RRTs collected nasopharyngeal (NP) and oropharyngeal (OP) swabs into a single tube and transported them to the KEMRI-Wellcome Trust Research Programme (KWTRP) for SARS-CoV-2 diagnosis. The laboratory diagnostic protocol for SARS-CoV-2 at KWTRP has been described elsewhere^8,10^ Briefly, the positive infections were identified using a two stage procedure, first, viral RNA purification using a QIAGEN extraction kit followed by real-time reverse-transcription PCR (RT-PCR) using one or two of four SARS-CoV-2 detection assays that we deployed at the centre since the beginning of the pandemic namely the Berlin Charite protocol, Europe Virus Archive Global (EVA-g) protocol, Beijing Genomics Institute (BGI) protocol and Da An Commercial Kit protocol (Said et al., 2020).

### SARS-CoV-2 genome amplification, library preparation and sequencing

SAR-CoV-2 positive samples with a cycle threshold (Ct) of <35.0 were processed for whole genome sequencing using the ARTIC nCoV-2019 sequencing V.1 protocol (Quick, 2020). Viral RNA was extracted from 140 μl of NP/OP swabs using the QIAamp Viral RNA Mini Kit (Qiagen, Manchester, UK) according to the manufacturer’s instructions except that carrier RNA was excluded at the lysis step. The purified viral RNA was then titrated based on a dilution factor that was determined from the relative amount of virus material in a sample as determined by Ct score.

A reverse transcription (RT-PCR) reaction was carried out using the multiplex ARTIC primer-pools A and B. The reaction volumes was carried out at half the recommended amount from the ARTIC sequencing protocol V.1 (Quick, 2020) and the PCR thermocycling reactions were run for 40 cycles. Primer pool A and B PCR products were pooled together to make a total of 25 μl and cleaned using 1X AMPure XP beads (Beckman Coulter). The pellet was resuspended in 15 μl nuclease-free water and 1uL of the eluted sample was quantified using the Qubit dsDNA HS Assay Kit (ThermoFisher).

End-prep reaction was performed according to the ARTIC nCoV-2019 sequencing protocol with 200 fmol (50 ng) of cDNA and the NEBNext Ultra II End repair/dA-tailing kits and incubated at 20°C for 5 minutes and 65°C for 5 minutes. From this, 1.5μl of DNA after End-Prep was used for native barcode ligation using NEBNext Ultra II Ligation (E7595L). In the absence of the Ligation Module, this step was performed using 10.5μl of Blunt/TA Ligase Master Mix (M0367L). Incubation was performed at 20°C for 20 minutes and at 65°C for 10 minutes. All barcoded samples were pooled together. The pooled and barcoded DNA samples were cleaned using 0.4X AMPure XP beads followed by two ethanol (80%) washes and eluted in 35uL of nuclease free water. Adapter ligation was performed using 50ng of the pooled sample, NEBNext Quick Ligation Module reagents (E6056L) and Adapter Mix II (ONT) and incubated at room temperature for 20 minutes. Final clean-up was performed using 0.4X AMPure XP beads and 12 μl of Short Fragment Buffer (ONT). The library was eluted in 15μl Elution Buffer. The final library was normalized to 15ng prior to loading on a flow-cell and sequencing with a MinION Mk1B device.

### Genome assembly

FAST5 files were base-called and demultiplexed using Guppy v4.0.11. Barcode indexes and sequencing primers were removed prior to consensus assembly of the sequences by aligning reads from each sample to reference SAR-CoV-2 genome (Genbank accession MN908947.3) as described in the ARTIC bioinformatics protocol. We modified the default subsampling from 200 to 5000 reads. Consensus sequence positions with insufficient (coverage <10 reads) genome coverage were masked with ambiguous bases (N). A basic quality control of the sequences was conducted to remove sequences with 20% or more ambiguous nucleotide bases (>4,000) from the analysis.

### SARS-CoV-2 lineage and clade assignment

**S**equences were assigned putative dynamic lineages (reference) using the Pangolin toolkit (version 2.0.4) with pangoLEARN (version 2020-07-20) and SARS-CoV-2 major clades were assigned through a locally installed instance of NextClade (release-0.2.4) (https://github.com/neherlab/webclades).

### Sampling from global dataset

A total of 62,644 SARS-CoV-2 whole genome sequences were downloaded from the Global Initiative on Sharing All Influenza Data (GISAID) sequence database (https://www.gisaid.org/) at a single time point. The sequences were filtered to remove erroneous, incomplete and improperly formatted data. Sequences shorter than 29,500 nucleotides those that were not accompanied by the day and month of collection were excluded. All sequences collected later than 24^th^ June 2020 were also excluded. The resulting dataset (n=) was then grouped based region, country and pangolin lineage and two sequences were picked to represent each lineage from each country. The remaining 1,439 sequences were then subjected to a quality check based on NextClade quality filters and visual inspection. The sequences that passed this quality filter (n=983) were referred to as the global dataset and were used to place the Coastal Kenya sequences within a global phylogenetic context.

### Maximum likelihood phylogenetic analysis

The set of sub-sampled global sequences (n=983) obtained from GSAID were combined with 274 sequences from Kenyan coast and aligned using MAFTT v7.310 (refs). The alignment file was manually inspected, the first and last 30 characters were masked including any base within a 7-base-pair window with more than two uninformative bases. Remaining ambiguous bases in the 3; and 5 UTR regions were simply removed from the alignment. Known problematic regions (ref) including those with more than 0.5 ambiguous or missing information were automatically masked. To place SARS-CoV-2 sequences collected from the coast within a global content, model selection was conducted and inferred using IQTREE (version 2.0.3) model selection tool and a maximum likelihood phylogenetic tree were estimated using the GTR+G+R3 substitution model. To estimate evolutionary relationships between sequences collected in coastal Kenya, a separate maximum likelihood phylogenetic tree was inferred from an alignment of 274 coastal Kenya sequences in addition to the NC_045512.2 reference sequence using RAxML-NGS v0.9.0) and run with 1000 bootstraps. The tree was rooted using the reference sequence as an outgroup. We estimated the minimum number of importations by triangulating between the known number of travel cases and manual inspection of the phylogenetic tree.

### Estimation of a Bayesian time-scaled phylogeny

We used BEAST version 1.10.4 Bayesian phylogenetic framework to reconstruct a time-scaled phylogeny, estimate divergence time and the rate of nucleotide substitution from 274 sequences from the Kenyan coast. The BEAST analysis was parameterized with an uncorrelated log-normal relaxed-clock (UCLN) model with an exponential growth coalescent prior and the HKY85 substitution model. Four independent Markov chain Monte Carlo (MCMC) runs of 100 million generations were performed with sampling every 10,000 generations. MCMC convergence and log files from the runs were inspected using Tracer v1.7. A maximum clade credibility (MCC) tree was generated using TreeAnnotator tool.

## QUANTIFICATION AND STATISTICAL ANALYSIS

Statistical analysis was performed using R version 4.0.2 and are described in the figure legends and in the method details.

## Acknowledgements

The authors would like to thank members of the county rapid response teams who collected nasal and oral swabs, and all the frontline workers at the health and testing facilities. Special thanks to the COVID-19 testing team at KWTRP who conducted the laboratory PCR assays.

We are grateful to those who have shared genome sequence data on GISAID (https://www.gisaid.org/). We thank the ARTIC Network (https://artic.network/) for providing us with primers and technical input and teams from the Oxford Nanopore Technology for the technical and material support. We would also like to acknowledge Prof La Scola Bernard for providing us with a heat-inactivated SARS-CoV-2 positive control that were used to validate our sequencing assays.

This work was supported by the National Institute for Health Research (NIHR) (project references 17/63/82 and 16/136/33 using UK aid from the UK Government to support global health research, The UK Foreign, Commonwealth and Development Office and Wellcome Trust (grant# 102975; 220985)

G.G. is funded and supported by NIHR funded GeMVi and TIBA projects (Grant number 17/63/82 and 16/136/33). C.A. is funded by the Initiative to Develop African Research Leaders (IDeAL) through the DELTAS Africa Initiative [DEL-15-003]. The DELTAS Africa Initiative is an independent funding scheme of the African Academy of Sciences (AAS)’s Alliance for Accelerating Excellence in Science in Africa (AESA) and supported by the New Partnership for Africa’s Development Planning and Coordinating Agency (NEPAD Agency) with funding from the Wellcome Trust [107769/Z/10/Z] and the UK government. The views expressed in this publication are those of the authors and not necessarily those of AAS, NEPAD Agency, Wellcome Trust NIHR or the Department of Health and Social Care or the UK government.

This was submitted for publication with permission from Director of KEMRI.

## Author Contributions

G.G., D.J.N., C.A., I.O., G.W., S.M.K, A.A., P.B. Conceptualised and designed the study G.G., S.K.M., Z.R.L., D.O.O., J.M.M. performed sequencing and provided laboratory support C.A., G.G., D.J.N. Provided phylogenetic analysis

B.T., I.O., A.A., E.B., C.A., K.K., R.A., J.K., E.M., B.K., T.S., M.M., J.N., J.O., B.S., provided administration and support for sample collection

E.O., G.G., C.A., S.K.M., provided data curation and analysis

G.G., C.A., D.J.N., G.W., I.O., B.T., P.B., wrote the original manuscript draft All contributors wrote, reviewed and edited the final manuscript

---

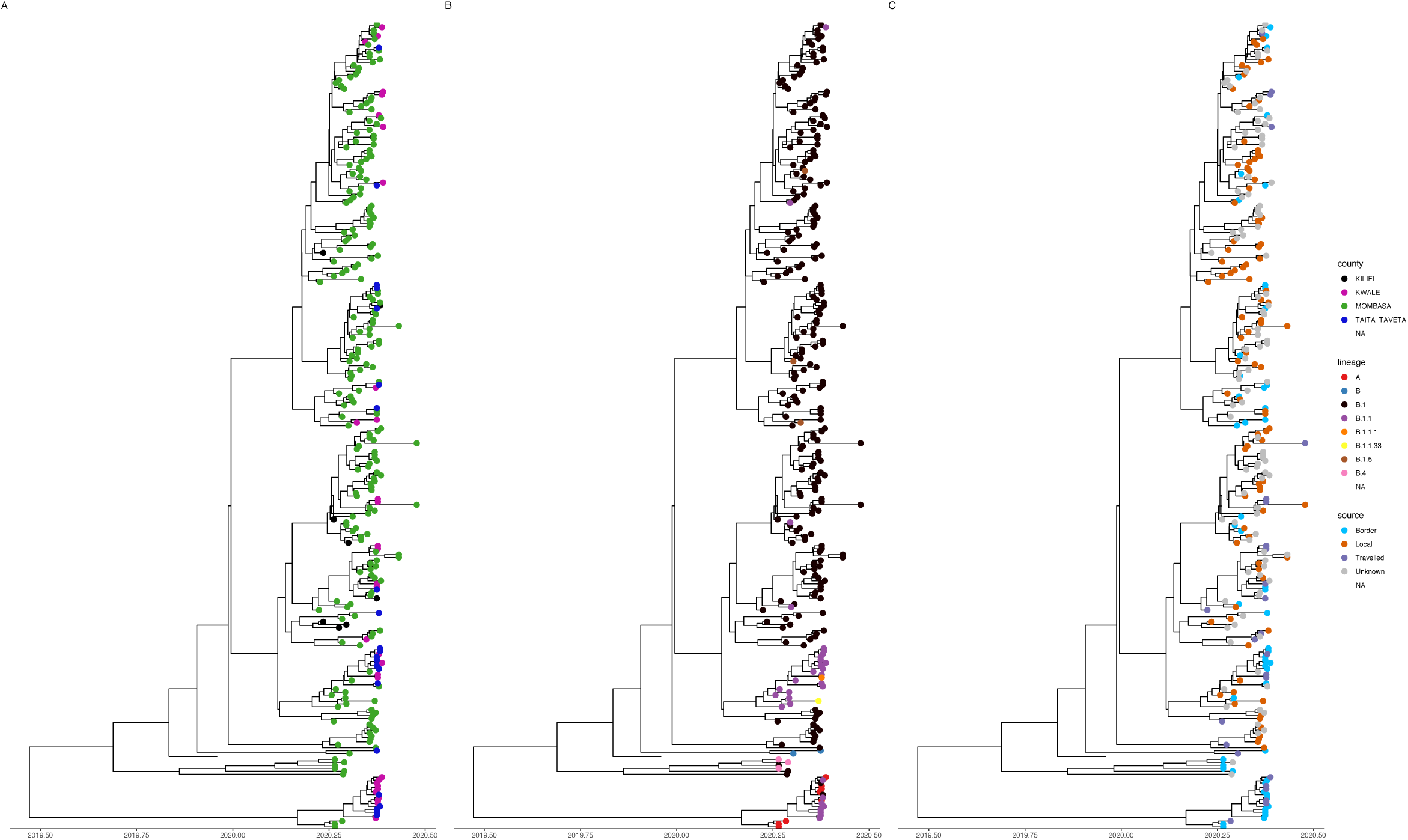

