## Supplementary figures and images for "Tracking the introduction and spread of SARS-CoV-2 in coastal Kenya"

### supplementary figure 1

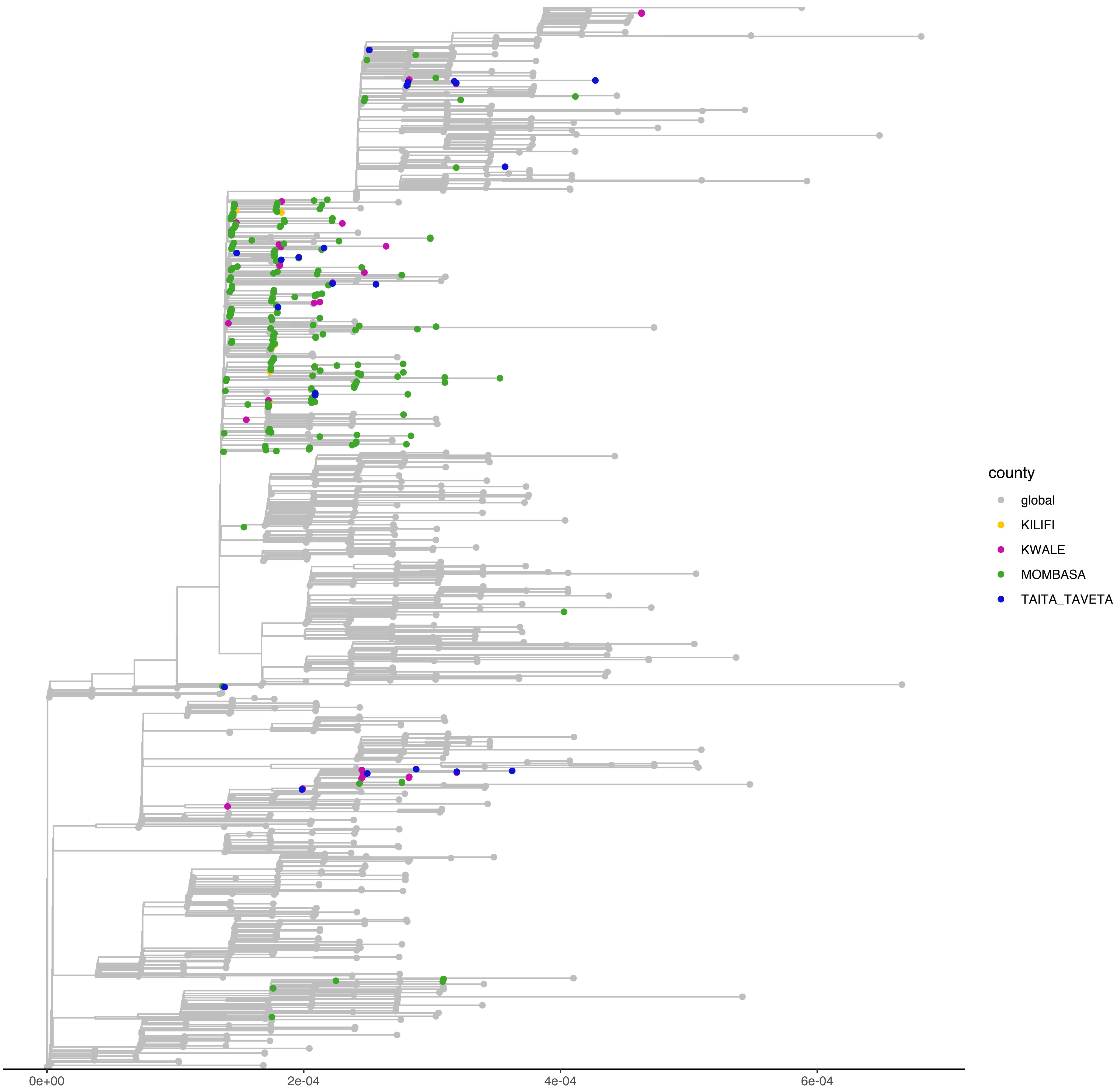

### supplementary figure 2

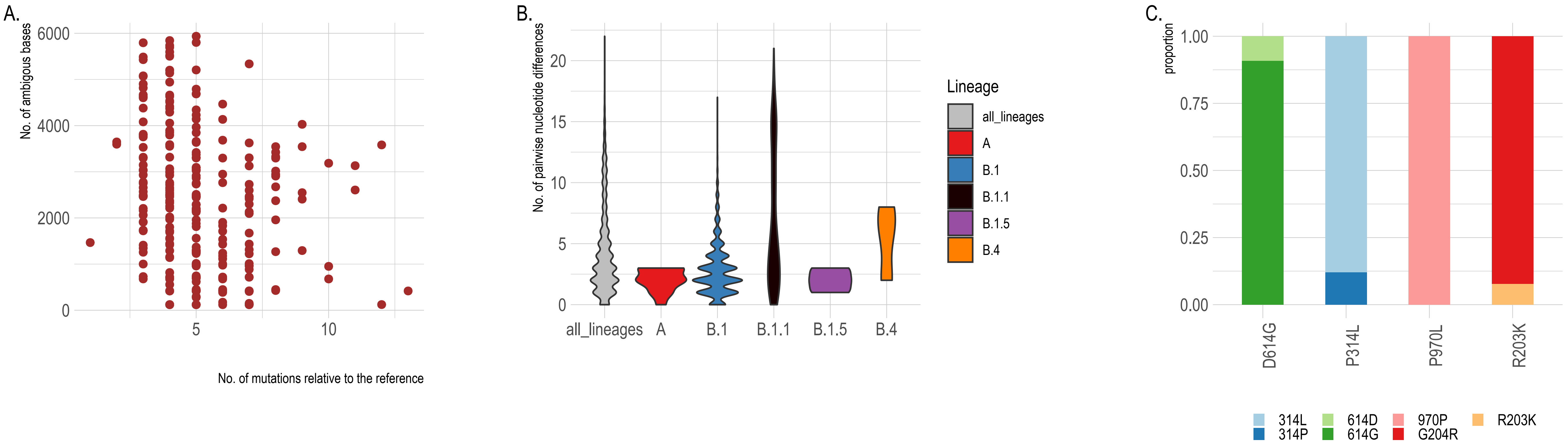

### supplementary figure 3

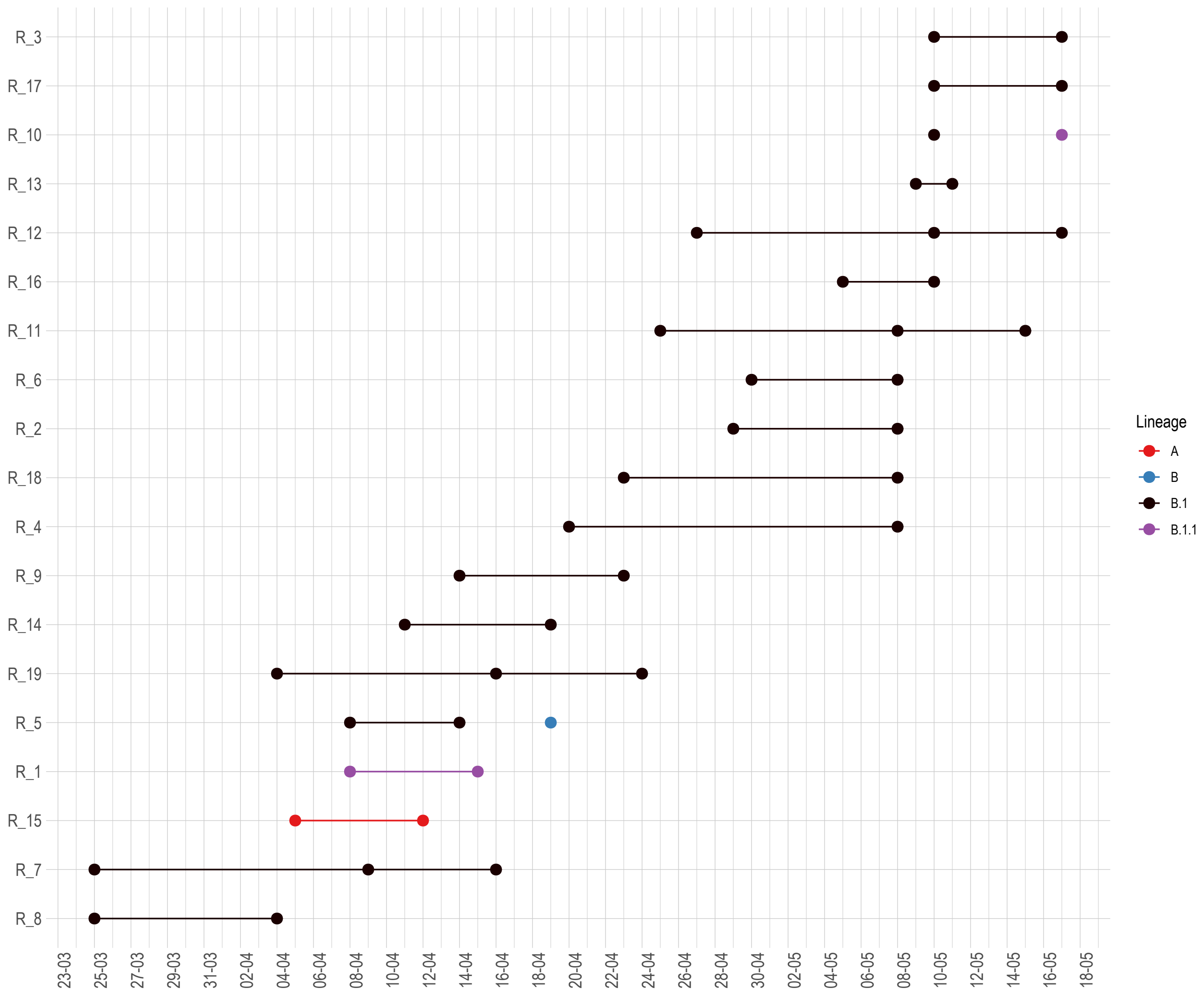

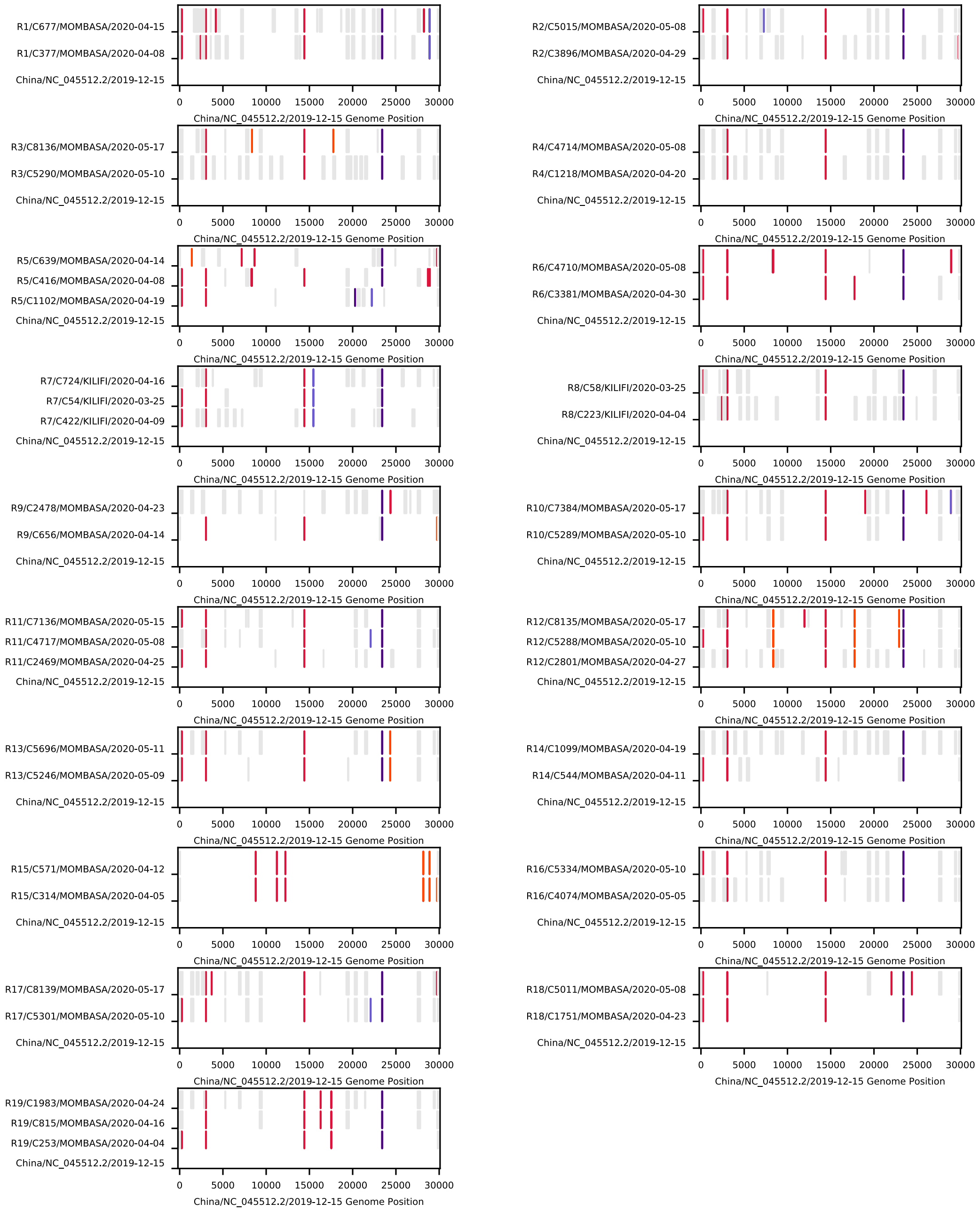
