## supplementary table 1 and 2 for "Tracking the introduction and spread of SARS-CoV-2 in coastal Kenya"

|  | **Kilifi (N=8)** | **Kwale (N=32)** | **Mombasa (N=212)** | **Taita Taveta (N=22)** | **Total (N=274)** |
| --- | --- | --- | --- | --- | --- |
| **Sex** |  |  |  |  |  |
| Female | 4 (50.0%) | 2 (6.2%) | 75 (35.4%) | 0 (0%) | 81 (29.6%) |
| Male | 2 (25.0%) | 29 (90.6%) | 118 (55.7%) | 20 (90.9%) | 169 (61.7%) |
| Unknown | 2 (25.0%) | 1 (3.1%) | 19 (9.0%) | 2 (9.1%) | 24 (8.8%) |
| **Age** |  |  |  |  |  |
| Mean (SD) | 37.0 (11.8) | 35.3 (12.3) | 41.4 (16.5) | 38.2 (8.82) | 40.3 (15.6) |
| Median [Min, Max] | 35.0 [30.0, 63.0] | 33.5 [0, 56.0] | 40.0 [1.00, 85.0] | 37.0 [22.0, 56.0] | 39.0 [0, 85.0] |
| Missing | 1 (12.5%) | 0 (0%) | 10 (4.7%) | 0 (0%) | 11 (4.0%) |
| **Age category** |  |  |  |  |  |
| 0-9 | 0 (0%) | 1 (3.1%) | 5 (2.4%) | 0 (0%) | 6 (2.2%) |
| 10-19 | 0 (0%) | 1 (3.1%) | 9 (4.2%) | 0 (0%) | 10 (3.6%) |
| 20-29 | 0 (0%) | 8 (25.0%) | 34 (16.0%) | 3 (13.6%) | 45 (16.4%) |
| 30-39 | 6 (75.0%) | 12 (37.5%) | 50 (23.6%) | 9 (40.9%) | 77 (28.1%) |
| 40-49 | 0 (0%) | 4 (12.5%) | 35 (16.5%) | 7 (31.8%) | 46 (16.8%) |
| 50-59 | 0 (0%) | 6 (18.8%) | 39 (18.4%) | 3 (13.6%) | 48 (17.5%) |
| 60-69 | 1 (12.5%) | 0 (0%) | 21 (9.9%) | 0 (0%) | 22 (8.0%) |
| 70-79 | 0 (0%) | 0 (0%) | 6 (2.8%) | 0 (0%) | 6 (2.2%) |
| 80-89 | 0 (0%) | 0 (0%) | 3 (1.4%) | 0 (0%) | 3 (1.1%) |
| Unknown | 1 (12.5%) | 0 (0%) | 10 (4.7%) | 0 (0%) | 11 (4.0%) |
| **Case history** |  |  |  |  |  |
| Local | 2 (25.0%) | 1 (3.1%) | 90 (42.5%) | 0 (0%) | 93 (33.9%) |
| Travel associated | 1 (12.5%) | 17 (53.1%) | 8 (3.8%) | 0 (0%) | 26 (9.5%) |
| Border | 0 (0%) | 13 (40.6%) | 18 (8.5%) | 22 (100%) | 53 (19.3%) |
| Unknown | 5 (62.5%) | 1 (3.1%) | 96 (45.3%) | 0 (0%) | 102 (37.2%) |
| **Symptoms** |  |  |  |  |  |
| Asymptomatic | 4 (50.0%) | 17 (53.1%) | 98 (46.2%) | 15 (68.2%) | 134 (48.9%) |
| Symptomatic | 3 (37.5%) | 0 (0%) | 41 (19.3%) | 1 (4.5%) | 45 (16.4%) |
| Unknown | 1 (12.5%) | 15 (46.9%) | 73 (34.4%) | 6 (27.3%) | 95 (34.7%) |

**Table S1:** Demographic characteristics of SARS-CoV-2 whole genome sequenced samples (>80% complete) collected between March and June 2020 (n=274) from coastal Kenya stratified by county. The case history demographic characteristic was derived from both self-reported travel history and presentation at a border point. Local case-history refers to individuals that did not report a history of travel and neither were they screened at a port of entry. Individuals whose case histories were not filled, or information was missing were labelled as unknown.

| Phase | Eligibility | Comments |
| --- | --- | --- |
| Phase 1 | - Showed specific symptoms of respiratory illness - Recent international travel - Close contacts of a confirmed case | Before confirmed case |
| Phase 2 | - Showed specific symptoms of respiratory illness - Recent international travel - Close contacts of a confirmed case - International visitors were taken to isolation centres | After first confirmed case |
| Phase 3 | - Mass testing rolled out at KPA and general public in Mombasa island | After suspected community transmission |
| Phase 4 | - Mass testing for truck drivers entering Kenya | A number of cases reported by Uganda border surveillance team |

**Table S2:** A table summarising the public health responses that were taken by the Ministry of Health in response to increasing number of cases. The definitions provided influenced the decision on who was tested. Here we divided the responses into 4 phases.
